# The vertical velocity step test and vertical sinusoidal harmonic acceleration test

**DOI:** 10.64898/2026.11.16.26368195

**Authors:** Fumihiro Mochizuki, Erin Williams, Madison Hawthorne, Bernat Miro, Alexander Kiderman, Michael Hoffer

## Abstract

**OBJECTIVE:** Current rotational chair testing assesses only horizontal semicircular canal (SCC) function. This study characterizes vertical SCC responses and provides preliminary descriptive reference ranges using sinusoidal harmonic acceleration (vSHA) and velocity step (vVST) paradigms.

**METHODS:** Forty-one healthy adults were tested with an adapted rotational chair to position the left anterior-right posterior (LARP) and right anterior-left posterior (RALP) SCCs in the Earth-horizontal plane. vSHA (at 0.08, 0.16, and 0.64 Hz) and vVST (at 100⁰/s) were performed. Gain, phase, asymmetry, time constants (TC), and maximum slow-phase eye velocity (MSPEV) were compared across planes. Correlations with video head impulse test (vHIT) gain were also examined.

**RESULTS:** vSHA gain (LARP: 0.37-0.43; RALP: 0.38-0.42) was lower than horizontal gain (0.59-0.63), consistent with previously reported vertical canal measurements. Cross-plane Friedman tests confirmed significantly lower vertical than horizontal gain at all three frequencies (all p < .001). Vertical phase lead exceeded horizontal at 0.08 and 0.16 Hz (all p-adjusted ≤ .021). Vertical TC were significantly shorter than horizontal for both per- and post-rotation (all p-adjusted < .001), and per- and post-rotation TC did not differ significantly in any plane. No significant LARP-RALP differences or vHIT-vVST correlations were found.

**CONCLUSIONS:** vSHA and vVST characterize vertical SCC function non-redundantly with horizontal testing and vHIT. In the vertical planes, gain values and TCs were reduced, which represents a pattern consistent with reduced velocity storage. Validation in larger and clinical samples is required before clinical adoption.

## 1. Introduction

Rotational stimuli delivered across physiologically relevant frequencies (0.001-1.00 Hz) enables comprehensive assessment of the vestibulo-ocular reflex (VOR), offering insight into the integrity and dynamic range of the vestibular system ^1^, particularly the semicircular canals (SCCs) ^2, 3^. Unfortunately, current rotational testing paradigms only stimulate the horizontal semicircular canals via Earth-vertical axis rotation, leaving vertical SCC function unexamined. Further, while the video head impulse test (vHIT) can target vertical semicircular canal responses, it operates during brief, high-acceleration head impulses ^4^ and is limited by technical and practical challenges, including difficulty of administration and measurement variability ^5–7^.

Among the primary assessments in a standard rotational test battery, the velocity step test (VST) and sinusoidal harmonic acceleration (SHA) test offer complementary assessments of VOR dynamics. The VST is useful for both identifying unilateral SCC hypofunction and distinguishing centrally mediated conditions such as vestibular migraine and motion sickness ^8, 9^. Two primary response metrics are reported for the VST: the time constant (TC), i.e., the time it takes for the rotation-induced VOR velocity to decline to 37% of its peak, and the maximum slow-phase eye velocity (MSPEV). These are classically measured across four conditions, per-rotation and post-rotation in the rightward clockwise (CW) and leftward counterclockwise (CCW) directions ^10^. In vestibular migraine, for example, prolonged TC have been observed across multiple VST conditions ^8^.

In healthy individuals, VST responses are expected to exhibit a high degree of symmetry between rightward and leftward rotations. This balance reflects equal excitatory and inhibitory drive between paired semicircular canals, consistent with Ewald’s laws of canal function. Deviations from this symmetry, such as direction-specific prolongation of TC or reduced MSPEV, may indicate unilateral canal weakness or central asymmetry in velocity storage mechanisms. Evaluating the symmetry of responses thus provides an intra-individual control and reference for identifying pathological deviations across both horizontal and vertical canal planes.

In contrast, the SHA test delivers a smooth and continuous oscillatory vestibular stimulus across low to middle range frequencies. SHA allows for insight into VOR gain and phase, making it useful in detecting frequency-dependent vestibular deficits that may be missed by more abrupt or high-velocity stimuli. Phase refers to the timing relationship between eye movement and head movement. Increased phase lead is often noted in subjects with vestibular pathology, particularly those with peripheral vestibular lesions ^11, 12^.

Though recent advancements have enabled the adaptation of existing paradigms for vertical SCC assessment ^13, 14^, clinical investigation in this area remains limited. To date, a single study conducted by Morita et al. has examined vertical canal responses during VST, demonstrating reduced MSPEV and TC in the vertical SCCs relative to the horizontal SCCs. Therefore, in this work, we sought to characterize responsivity of the vestibular system following two distinct velocity-profile stimuli. These stimuli differ both in their kinematic properties and their capacity to probe semicircular canal function; VST provides insight into velocity storage in response to abrupt accelerations, while SHA yields a frequency-dependent profile of VOR gain and phase during sustained oscillatory stimuli.

Together, vertical VST (vVST) and vertical SHA (vSHA) extend the application of rotational stimuli to the veritcal SCC planes (i.e., left anterior/right posterior [LARP] and right anterior/left posterior [RALP]), building on the foundational work of Morita et al. ^15^ with the potential to complement current clinical diagnostic tools for central and peripheral vestibular disorders.

## 2. Methods

### 2.1. Study Participants

Overall, we enrolled 41 participants (18 males, 23 females; aged 18 to 51 years [mean age 26.3 ± 6.7 years]) with no history of vestibular disorders or traumatic brain injury. All participants provided written informed consent before being enrolled in the study. This study was approved by the Institutional Review Board at the University of Miami (#20190034). Among the 41 enrolled participants, 27 completed vVST testing across all three canal planes, and 24 completed the full protocol (both vSHA and vVST in all three planes). The remaining 14 participants were enrolled before the vertical testing protocol had been finalized and were not retested (**Figure 1**).

**Figure 1.**
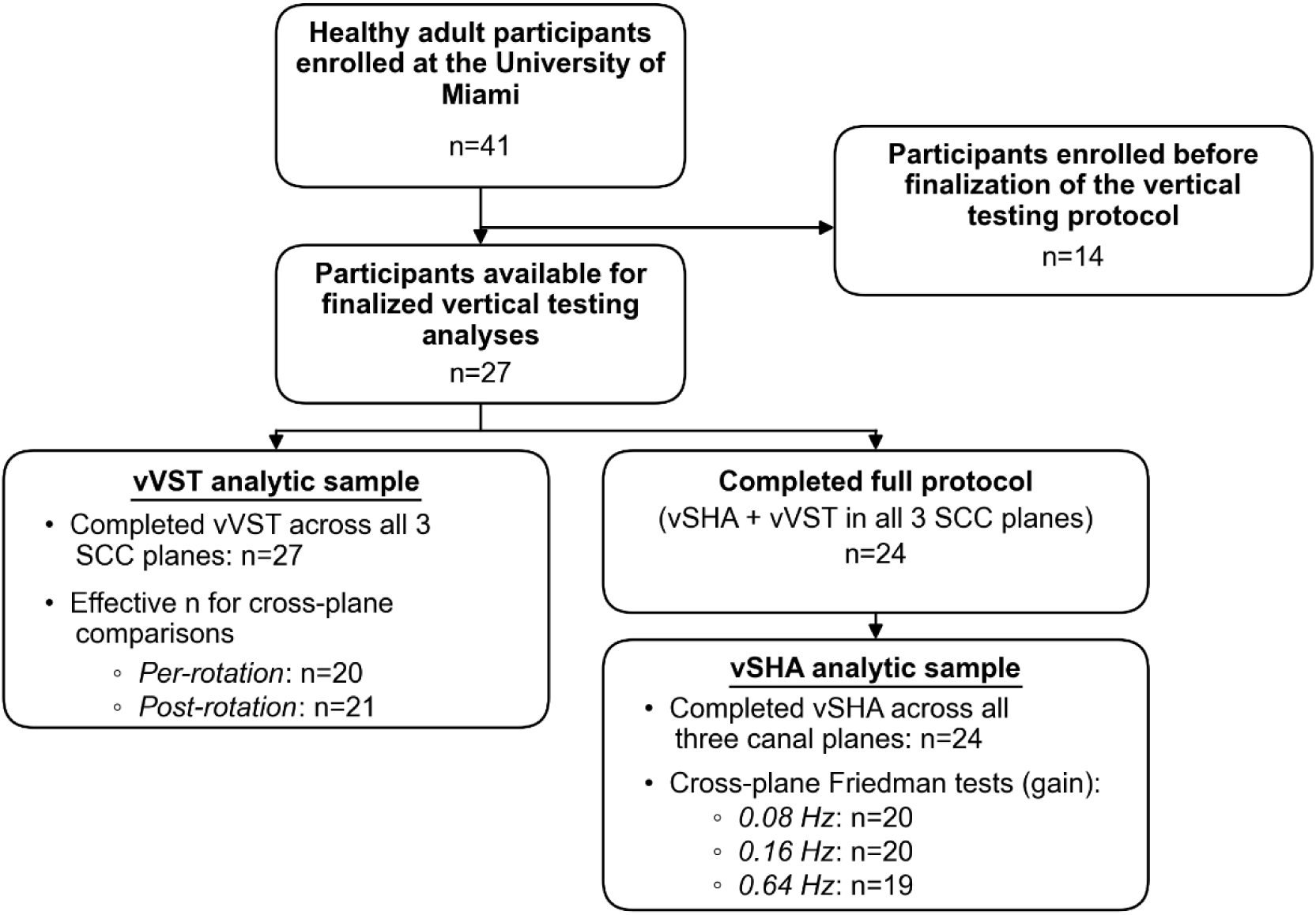
Participant flow and analytic sample sizes for vertical sinusoidal harmonic acceleration (vSHA) and vertical velocity step testing (vVST). Of 41 enrolled participants, 27 completed vVST testing across all three canal planes, and 24 of these completed the full protocol, including both vSHA and vVST across all three planes. The remaining 14 participants were enrolled before the vertical testing protocol was finalized and were not retested. Effective sample sizes for cross-plane analyses varied according to data completeness for each frequency and rotation condition.

### 2.2. Device

Whole-body rotational tests were delivered using the Neuro-Otologic Testing Center (NOTC) device supplied by Spryson America, Inc. (Pittsburgh, PA). Eye position was measured using binocular infrared video-oculography with a spatial resolution of 0.1 degrees horizontal and vertical at a frame rate of 250 frames per second. The video-oculography goggles included an embedded 6-degrees-of-freedom sensor comprising a 3-axis angular rate sensor and a 3-axis linear accelerometer.

The back of the chair was modified to manually tilt backwards 70 degrees, with an additional customizable headrest for LARP and RALP adjustment by rotation of the headrest ± 45 degrees to the left or right (**Figure 2**) to orient the vertical SCC pairs in the earth-horizontal plane. The rotational chair was enclosed in a light-proof enclosure, and participants were secured with a four-point restraint system with the head stabilized against the headrest.

**Figure 2.**
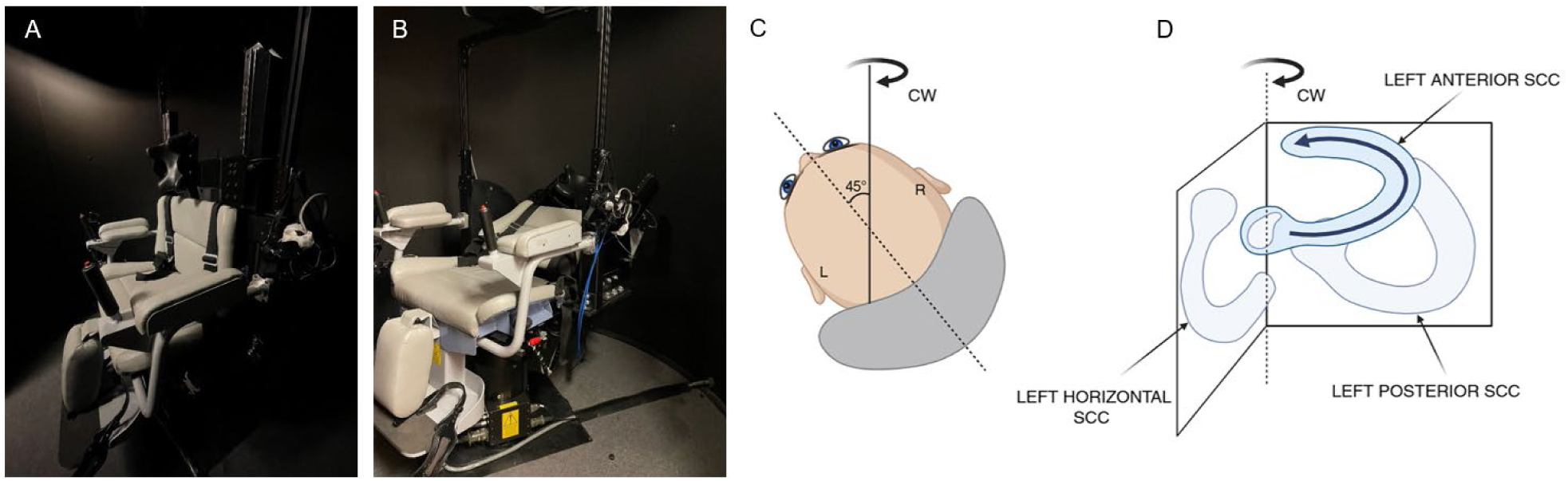
Vertical semicircular canal testing setup using the Neuro-Otologic Testing Center (NOTC) rotational chair device [Spryson America, Inc., Pittsburgh, PA]. (A) Rotational chair in the upright position for horizontal semicircular canal testing. (B) Chair pitched 70 degrees from vertical to position the vertical semicircular canal planes. (C) Subject’s head yawed 45 degrees to the right to align the left anterior-right posterior (LARP) canal plane with the Earth-horizontal plane (viewed from behind). (D) Schematic of the left semicircular canals during rightward (**CW**) rotation in the LARP position, illustrating the direction of ampullofugal endolymph flow in the left anterior canal.

#### 2.2.1. Procedure

First, each participant was seated into the rotational chair and securely restrained with the head pitched down such that horizontal SCC plane was aligned with the Earth-horizontal plane. The head was positioned so that the axis of rotation was approximately 2 cm in front of the interaural line. The video-oculography goggles were calibrated using fixation targets at +10 degrees horizontally and +10 degrees vertically, followed by a standard oculomotor testing battery to rule out central pathology. Horizontal vestibular assessments were conducted, including the computerized rotational head impulse test, SHA at 0.08, 0.16, and 0.64 Hz, and VST at 125⁰/s². Head alignment with the Earth-horizontal plane was confirmed by visual inspection of the Reid’s plane (a line from the inferior orbital rim to the center of the external auditory canal).

For assessment of the vertical canals, the chair was manually tilted back 70 degrees with the headrest rotated 45 degrees to the left or right. By adjusting the headrest position, we were able to orient the vertical SCC plane being tested parallel to Earth-horizontal (**Figure 2**). Then, a separate calibration was performed, where a laser located at the center of the goggles was used to locate a stationary light-emitting diode (LED) target in the primary position attached to the interior wall of the enclosure, approximately 1m away from the rotational chair. Eye-velocity calibration was performed by rotating the participant sinusoidally about an Earth-vertical axis at a frequency of 0.25 Hz with an amplitude of ±10 degrees while the earth-fixed LED target remained illuminated and the participant looked at the target. After the calibration, each participant underwent vSHA at 0.08, 0.16, and 0.64 Hz and vVST in each vertical SCC plane (**Figure 3** demonstrates LARP positioning). This calibration procedure established a known relationship between ocular angular displacement and video-oculography pixel output for each participant in the tilted position, ensuring that gain calculations were referenced to calibrated eye-velocity signals. For the duration of the assessment, the participants were given verbal tasks to maintain alertness.

**Figure 3.**
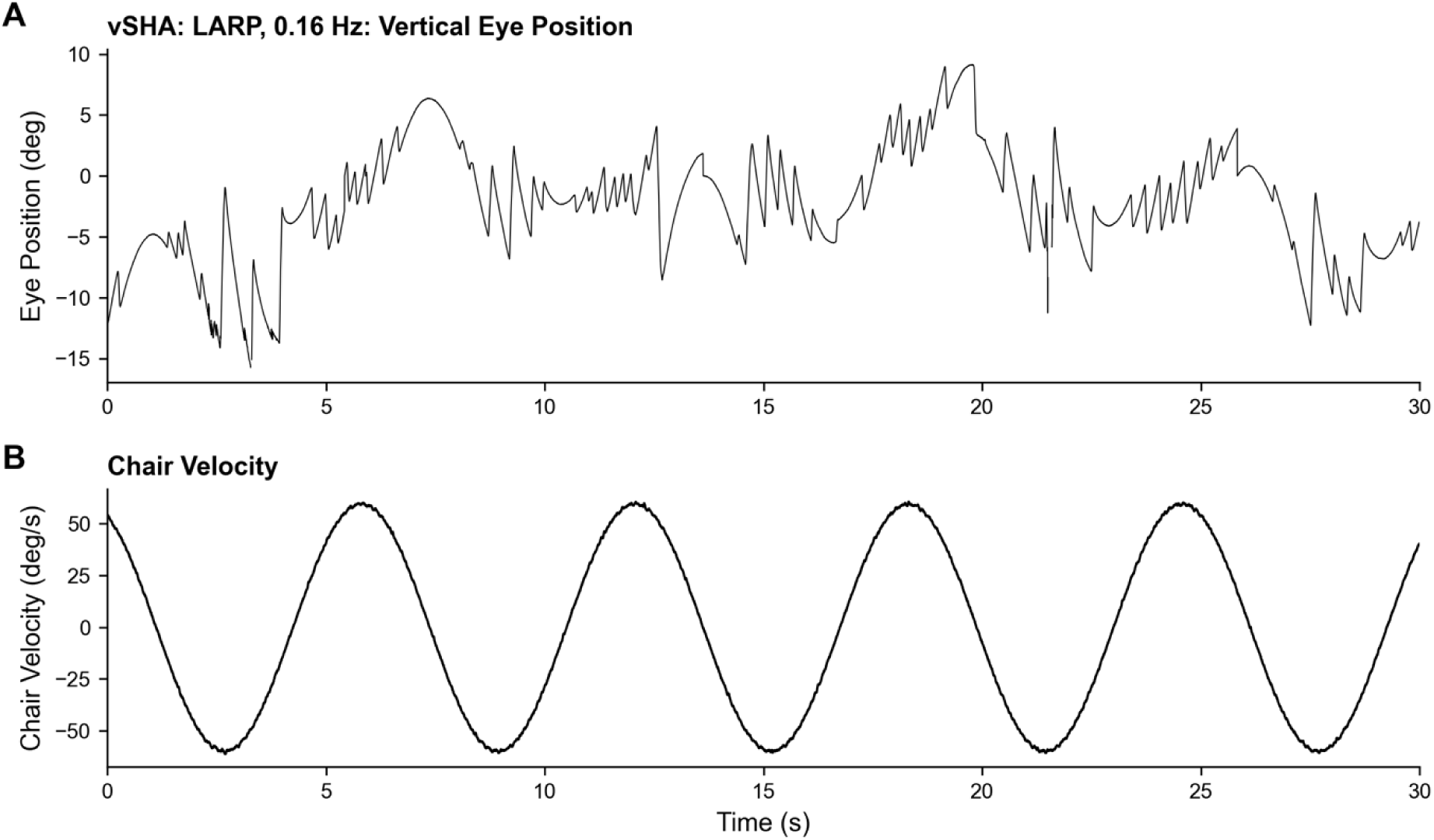
Representative nystagmus recording during vSHA testing at 0.16 Hz, illustrating bilateral stimulation of the vertical semicircular canals. The rotational chair follows a sinusoidal velocity profile, gradually accelerating until reaching peak velocity, then decelerating to a stop before reversing direction. (A) Alternating up-beating and down-beating vertical nystagmus recorded with the head tilted 45 degrees to the right for left anterior/right posterior (LARP) canal plane positioning. (B) Sinusoidal yaw-axis velocity profile of the rotational chair. Eye position is plotted in degrees (positive values = upward); chair velocity is plotted in degrees per second.

Throughout this manuscript, CW and CCW refer to the direction of chair rotation about the Earth-vertical axis as viewed from above. As with horizontal VST, vertical VST involved the participants undergoing acceleration at 125⁰/s² clockwise from 0⁰/s to 100⁰/s, rotation at this speed for 60 s, and then an abrupt deceleration at 125⁰/s² to held at zero speed for 60 s. This cycle was repeated in the counterclockwise direction. The order in which RALP and LARP were performed was randomized.

The primary excitatory SCC stimulated during each acceleration or deceleration was determined by Ewald’s first law based on the direction of endolymph flow relative to each canal’s ampulla. The excited canal for each of the 12 vVST conditions is listed in **Table 1**.

**Table 1.**
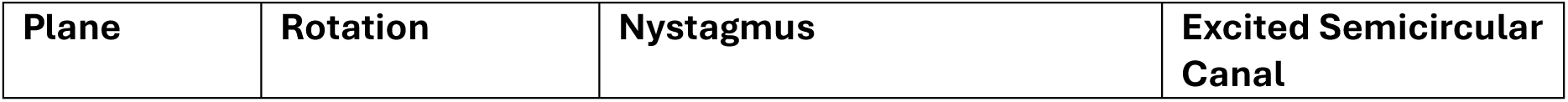
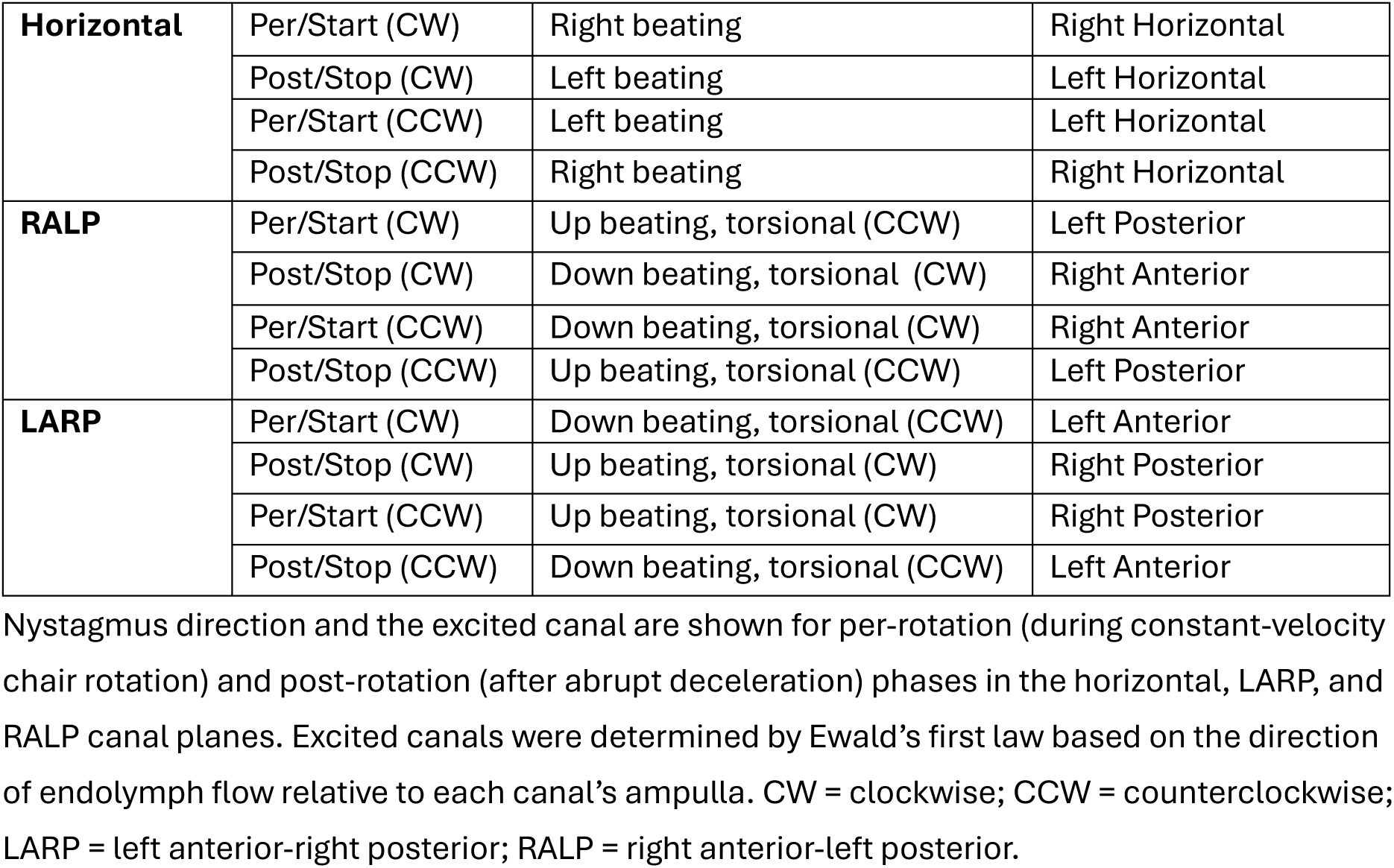
Induced nystagmus direction and primarily excited semicircular canal for each velocity step test (VST) condition.

#### 2.2.2. Data acquisition and processing

During each test, left and right eye position and head angular velocity were continuously collected at a sampling rate of 250 Hz. Horizontal eye movement was used when evaluating the horizontal SCCs and vertical eye movement for the vertical SCCs. To eliminate a small drive-train mechanical delay (∼28 ms) that would otherwise introduce frequency-dependent phase errors at higher stimulus frequencies, all stimulus reference trajectories were derived directly from actual head movement rather than the controller’s commanded chair signal. Because head tilt during vertical canal testing distributes physical rotation across all three gyroscope channels, principal component analysis (PCA) was used to identify the primary axis of rotation and project the 3-axis velocity vector onto this axis, reconstructing the full scalar angular velocity delivered to the head. Across all recordings, the first principal component captured a mean of 99.8% of the gyroscope variance (range: 97.5-100.0%), confirming that the single-axis rotation assumption was well satisfied. Ǫuick-phase nystagmus segments were identified and removed using an adaptive multi-criterion detector that iteratively fits a least-squares sinusoid at the stimulus frequency and flags residual outliers, acceleration transients, and samples exceeding a 200⁰/s physiologic ceiling consistent with the saccadic main sequence ^16, 17^; flagged runs were dilated by ±32 ms (8 samples) to capture saccadic ramps. The velocity-threshold and derivative-based approach to separating slow and fast phases follows established nystagmus analysis methods ^18, 19^.

#### 2.2.3. vHIT

vHIT was performed using the ICS Impulse system (Otometrics, Denmark). Participants were seated and asked to fixate a target at approximately 1.5m while the examiner delivered brief, unpredictable head rotations of 10-20° in each SCC plane (≥15 impulses per direction). Gain was computed as area-under-the-curve gain by the Otometrics system, excluding covert saccades ^20, 21^. Catch-up saccades were classified as pathological using previously published criteria ^21, 22^. The vHIT data reported herein were collected as part of the same testing session and include overlapping participants with those reported in Williams et al. ^13^; however, the present analysis focuses on the correlation between vHIT gain and rotational chair parameters, which was not examined in the prior study.

### 2.3. Vertical Tests

#### 2.3.1. Vertical sinusoidal harmonic acceleration test (vSHA)

For each cycle, gain was computed per half-cycle, with halves delimited by zero-crossings of the PCA-reconstructed head angular-velocity signal. Within each half, peak slow-phase eye velocity (SPV) was estimated by sliding a frequency-scaled window across slow-phase intervals and fitting a linear regression of eye position versus time. Per-half gain was calculated as peak SPV divided by the amplitude of a least-squares sinusoid fitted at the stimulus frequency to the PCA-reconstructed head angular velocity, and cycle-level gain was computed as the mean of the two per-half gains. As a cross-check, gain was also computed as the amplitude of a least-squares sinusoidal fit of eye velocity, fitted to the samples retained after quick-phase removal, divided by the same head-velocity amplitude. Phase was computed from the sinusoidal fit as the temporal lag between slow-phase eye velocity and the gyroscope-reconstructed velocity (0° = perfectly compensatory; positive = lead). Directional asymmetry was computed from each subject’s mean per-half values across accepted half-cycles as 100 *x* ((*peak SPV*_*H*1 − *peak SPV*_*H*2))/((*peak SPV*_*H*1 + *peak SPV*_*H*2)). This metric uses peak slow-phase velocity per half-cycle rather than the sinusoidal fit, and is therefore independent of the fit-quality gate.

Data were filtered through three a priori quality gates: half-cycles with >55% non-slow-phase samples were excluded, phase summaries excluded cycles with sinusoidal fit R² < 0.50 or phase outside ±60°, and observation-level estimates were excluded if gain fell outside 0.05-1.2, phase exceeded ±60°, or spectral purity dropped below 60%. Final cycle inclusion was determined by manual review. After automated gating, one author (E.W.), not blinded to canal plane or frequency, inspected each cycle in an interactive viewer displaying raw and desaccaded eye velocity, PCA-reconstructed head angular velocity, and the sinusoidal-fit overlay. A cycle was retained only if it exhibited visibly clean slow phases; contaminated segments were removed at the half-cycle level, so a cycle could contribute a single surviving half (consistent with the automated data-loss gate). Half-cycle inclusion followed the reviewer’s decisions: a half-cycle accepted or rejected in review was included or excluded accordingly, a cycle accepted in review contributed both halves, and the automated per-half fit criterion applied only to half-cycles without a review decision. The retention figures and all SHA parameters reflect this combined automated-plus-manual screening.

#### 2.3.2. vVST

For vVST, outcome measures were the TC and the MSPEV. TC is defined as the time, in seconds, for the VOR response to decline to 37% of its maximum value. In VST, data were acquired during rotation in each direction (per-rotation) and after rotation stopped (post-rotation); thus, in total, there were four conditions for each plane. The four conditions were as follows: (1) during CW acceleration (per-CW), (2) after abruptly stopping CW rotation (post-CW), (3) during CCW acceleration (per-CCW), and (4) after abruptly stopping CCW rotation (post-CCW). The VST was performed in three planes: Horizontal, RALP, and LARP for a total of 12 conditions. **Figure 4** shows the primarily stimulated SCCs and the direction of the induced nystagmus in these 12 conditions. Only vertical eye movement was used for the calculation of MSPEV in the vertical SCC planes. Finally, to mitigate confusion, the name of the SCC on the excited side was used to label vVST results for the purpose of this manuscript.

**Figure 4.**
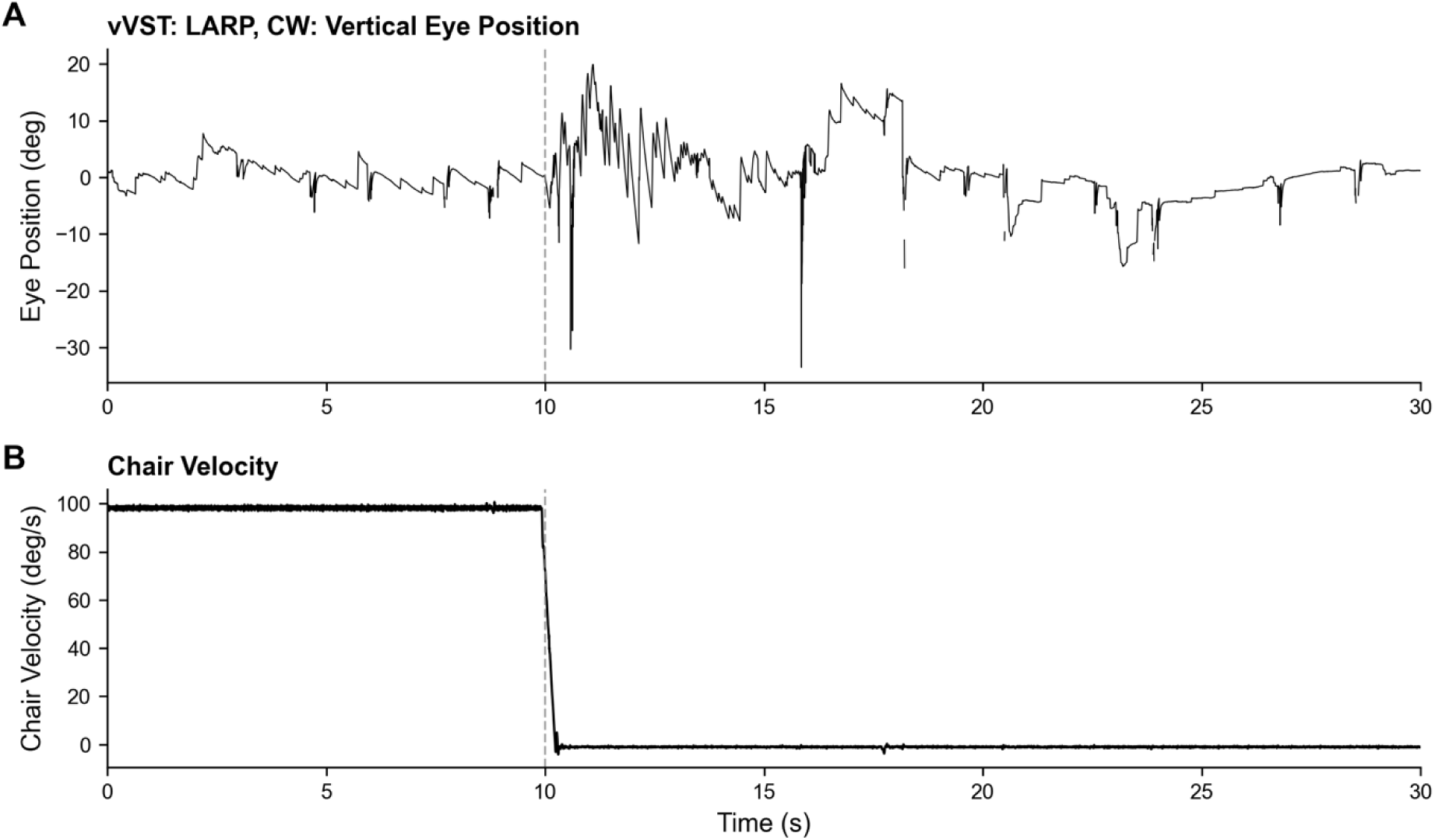
Eye position and rotational chair velocity during and after clockwise (CW) rotation with the subject in the left anterior-right posterior (LARP) position during the vertical velocity step test (vVST). (A) Vertical eye position (degrees; positive values = upward) over the course of the vVST stimulus, showing per-rotation nystagmus during constant-velocity rotation and post-rotation nystagmus following abrupt deceleration. (B) Chair angular velocity (⁰/s) during the per-rotation and post-rotation phases.

The chair angular velocity reference was reconstructed via PCA of the head-mounted gyroscope signal, as described for SHA. Per-rotation and post-rotation segments were identified automatically from the chair velocity profile. Ǫuick phases are distinguished from slow phases by their high acceleration ^18^ and, in nystagmus, beat in the direction opposite to the slow phase. We therefore removed samples whose velocity opposed the local slow phase (500 ms running median) by more than four times the local robust standard deviation of slow-phase velocity, with ±32 ms dilation to capture the onset and offset of each quick phase, together with samples exceeding 1.1 times the chair velocity in the compensatory direction or a 200⁰/s physiologic ceiling consistent with the saccadic main sequence ^16^ to exclude blink and tracking artifacts. MSPEV was measured as the maximum, in the compensatory direction, of a 500 ms rolling-median-smoothed slow-phase velocity trace over the first 5 seconds after segment onset, computed only where at least half of the samples in the window were retained ^15^. TC was measured directly, as the time from this peak until the smoothed slow-phase velocity fell to 37% of the peak and remained at or below that level for at least 1 s ^23, 24^. Segments were excluded if slow-phase data completeness was below 30%, MSPEV was below 5⁰/s or less than three times the robust variability of the smoothed trace over the final 10 s of the segment, or TC fell outside the 1–40 s plausibility window ^25, 26^.

### 2.4. Velocity storage index and canal pair asymmetry

A velocity storage index (VSI) was computed per canal plane as *VSI* = (*TC*_*post* − *TC*_*per*)/(*TC*_*post* + *TC*_*per*), where TC_post and TC_per are the post- and per-rotation time constants, respectively. Canal plane differences were assessed by Friedman tests with Benjamini-Hochberg-corrected pairwise Wilcoxon signed-rank tests. Canal pair asymmetry ratios (AR) were computed for per- and post-rotation conditions and compared with vHIT gain asymmetry using Spearman rank correlation.

### 2.5. Transfer function fitting

To estimate VOR time constants from the SHA gain-frequency relationship for cross-modality comparison with VST TCs, we attempted to fit a first-order high-pass transfer function *Gain*(*f*) = (*ωT*) / √(1 + (*ωT*)²), where *ω* = 2*πf* and *T* is the fitted time constant, to each subject’s three-frequency gain curve (0.08, 0.16, 0.64 Hz). Fits were performed using nonlinear least-squares (nlsLM, R package minpack.lm; T constrained to [1.0, 100] s). The intent was to derive an SHA-based TC for each canal plane that could be compared with the directly-measured VST TCs.

### 2.6. Statistical analysis

All descriptive statistics are reported as mean (± SD). Within-subject comparisons between canal planes were conducted using Wilcoxon signed-rank tests (two planes) or Friedman tests (three planes). Between-group comparisons, where applicable, used the Wilcoxon rank-sum test. Effect sizes for Wilcoxon signed-rank tests were calculated as the matched-pairs rank-biserial correlation, *r* = 4*V*/*n*(*n* + 1) − 1, where V is the sum of positive signed ranks and n is the number of non-zero paired differences. Absolute values are reported; direction is conveyed by the accompanying text. Cross-plane comparisons (three planes: horizontal, LARP, RALP) used Friedman tests with pairwise Wilcoxon signed-rank post-hoc tests. Within-plane two-group comparisons (LARP vs RALP across SHA frequencies; per-vs post-rotation within plane for VST) used paired Wilcoxon signed-rank tests. This unified nonparametric framework eliminates sphericity assumptions and ensures consistent effect-size reporting (Kendall’s W for omnibus, rank-biserial r for pairwise tests). Because cross-plane comparisons required complete data across the horizontal, LARP, and RALP planes, participants without complete data for a given frequency or rotation condition were excluded from that specific analysis; accordingly, analytic sample sizes varied across comparisons (**Figure 1**). For SHA asymmetry comparisons, absolute asymmetry values were used to avoid the sign artifact introduced by the H1/H2 canal-assignment convention (LARP and RALP code anterior/posterior canals in opposite H1/H2 slots). All p-values were corrected for multiple comparisons with the Benjamini-Hochberg false-discovery-rate procedure applied within each test family: the three pairwise comparisons following each significant Friedman omnibus; the LARP-RALP comparisons across the three SHA frequencies within each measure (gain, phase, absolute asymmetry); the six per-versus post-rotation comparisons (TC and MSPEV in each plane); the left-right canal-pair comparisons within each metric; the 24 vHIT-VST correlations; and the per-rotation asymmetry-ratio versus vHIT asymmetry correlations.

Preliminary descriptive reference data were derived for all SHA parameters (gain, phase, asymmetry) by plane and frequency and for all vVST parameters (TC, MSPEV) by canal and rotation direction. Descriptive statistics include the mean ± SD and the 5th-95th percentile range. Because the sample size (n = 16-26 per cell) is below the minimum recommended for definitive reference intervals (n ≥ 120; CLSI C28-A3), 90% bootstrap confidence intervals on the 5th and 95th percentile estimates are reported to characterize the precision of these preliminary limits.

## 3. Results

### 3.1. vSHA

For the horizontal plane (**Table 2**), gain was 0.60 at 0.08 Hz, 0.59 at 0.16 Hz, and 0.63 at 0.64 Hz. Phase values were near zero across all frequencies. For vertical canals, LARP gain ranged from 0.37 to 0.43 and RALP gain from 0.38 to 0.42 across the three frequencies. Within-subject Wilcoxon signed-rank tests comparing LARP and RALP gain at each frequency showed no significant differences after Benjamini-Hochberg correction (0.08 Hz: V = 113, p-adjusted = .945; 0.16 Hz: V = 93, p-adjusted = .945; 0.64 Hz: V = 142, p-adjusted = .184, r = 0.49). Absolute asymmetry (|asymmetry|) did not significantly differ between LARP and RALP at any frequency (0.08 Hz: V = 46, p-adjusted = .366; 0.16 Hz: V = 44, p-adjusted = .366; 0.64 Hz: V = 50, p-adjusted = .366), and phase did not differ between LARP and RALP at any frequency (all p-adjusted ≥ .427).

**Table 2.** Sinusoidal harmonic acceleration (SHA) gain, asymmetry (%), and phase (degrees) across the horizontal, LARP, and RALP semicircular canal planes at 0.08, 0.16, and 0.64 Hz.

| Plane | Horizontal |  |  | LARP |  |  | RALP |  |  |
|---|---|---|---|---|---|---|---|---|---|
| **Frequency (Hz)** | **0.08** | **0.16** | **0.64** | **0.08** | **0.16** | **0.64** | **0.08** | **0.16** | **0.64** |
| **Gain**<br>(Mean ± SD) | 0.60<br>(± 0.18) | 0.59<br>(± 0.17) | 0.63<br>(± 0.14) | 0.41<br>(± 0.10) | 0.37<br>(± 0.12) | 0.43<br>(± 0.10) | 0.42<br>(± 0.10) | 0.41<br>(± 0.14) | 0.38<br>(± 0.11) |
| 5th percentile | 0.34 | 0.31 | 0.39 | 0.28 | 0.17 | 0.29 | 0.27 | 0.25 | 0.22 |
| 95th percentile | 0.84 | 0.91 | 0.79 | 0.60 | 0.52 | 0.58 | 0.56 | 0.72 | 0.55 |
| **Phase**<br>(Mean ± SD) | 2.1<br>(± 4.1) | 1.4<br>(± 3.7) | 0.8<br>(± 3.9) | 11.1<br>(± 4.5) | 4.8<br>(± 5.9) | 1.2<br>(± 5.1) | 11.2<br>(± 4.9) | 5.2<br>(± 4.3) | 0.7<br>(± 5.6) |
| 5th percentile | -4.4 | -4.1 | -5.0 | 2.8 | -5.3 | -6.0 | 2.9 | -1.3 | -7.2 |
| 95th percentile | 7.2 | 4.1 | 6.9 | 15.8 | 11.6 | 9.3 | 16.3 | 13.0 | 9.0 |
| **Canal Asymmetry**<br>(Mean ± SD) | -4.6<br>(± 12.8) | 0.1<br>(± 10.7) | 1.7<br>(± 6.0) | -12.8<br>(± 15.8) | -1.8<br>(± 13.5) | 0.4<br>(± 11.9) | 13.0<br>(± 20.9) | 10.4<br>(± 21.3) | -1.8<br>(± 13.6) |
| 5th percentile | -22.5 | -20.0 | -6.8 | -32.8 | -25.3 | -16.2 | -13.9 | -15.4 | -17.6 |
| 95th percentile | 12.0 | 11.9 | 11.0 | 4.6 | 16.0 | 16.4 | 43.9 | 40.6 | 17.0 |

Values are presented as mean (± SD). Gain was calculated as the ratio of peak slow-phase eye velocity to PCA-reconstructed head angular velocity, averaged across the two half-cycles. Phase represents the temporal offset between stimulus and response; positive values indicate phase lead (eye velocity precedes chair velocity). Asymmetry (%) was calculated per subject as 100 ×((mean H1 gain − mean H2 gain))/ (mean H1 gain + mean H2 gain). LARP = left anterior-right posterior; RALP = right anterior-left posterior. Preliminary descriptive reference ranges (5th and 95th percentiles) are included for each measure.

#### 3.1.1. Three-plane comparison of SHA parameters

Gain differed significantly across planes at all three frequencies: 0.08 Hz (χ²(2) = 17.50, p < .001, Kendall’s W = 0.438), 0.16 Hz (χ²(2) = 19.20, p < .001, Kendall’s W = 0.480), and 0.64 Hz (χ²(2) = 26.00, p < .001, Kendall’s W = 0.684). Benjamini-Hochberg-corrected post-hoc Wilcoxon signed-rank tests confirmed that horizontal gain was significantly higher than both LARP and RALP at every frequency (all p-adjusted ≤ .001), while LARP and RALP did not differ at any frequency (all p-adjusted ≥ .061). Absolute asymmetry did not differ across planes at any frequency (0.08 Hz: χ²(2) = 0.14, p = .931; 0.16 Hz: χ²(2) = 2.53, p = .282; 0.64 Hz: χ²(2) = 4.62, p = .099). Phase differed across planes at 0.08 Hz (χ²(2) = 14.92, p < .001, Kendall’s W = 0.574) and 0.16 Hz (χ²(2) = 15.65, p < .001, Kendall’s W = 0.460) but not at 0.64 Hz (χ²(2) = 1.68, p = .431). Phase lead was greater in both vertical planes than in the horizontal plane at 0.08 Hz (LARP 11.1°, RALP 11.2°, horizontal 2.1°; both p-adjusted = .005, r = 0.93-1.00) and 0.16 Hz (LARP 4.8°, RALP 5.2°, horizontal 1.4°; both p-adjusted = .021, r = 0.69-0.78), with no LARP-RALP difference (p-adjusted ≥ .142).

### 3.2. vVST

Time constants (TC) were numerically longer in the post-rotation condition than during per-rotation in all three planes, and maximum slow-phase eye velocity (MSPEV) was numerically higher post-rotation in the horizontal and LARP planes (Table 3). Six paired Wilcoxon signed-rank tests (TC and MSPEV in each of three planes) were Benjamini-Hochberg corrected. After correction, no per-versus post-rotation difference was significant. For the horizontal plane, TC was 14.0 ± 3.8 s post-rotation versus 12.3 ± 4.8 s per-rotation (V = 242, raw p = .094, p-adjusted = .281, r = 0.38), and MSPEV did not differ (V = 246, raw p = .075, p-adjusted = .281, r = 0.40). LARP MSPEV (V = 171, p-adjusted = .306, r = 0.35) and the remaining vertical comparisons (all p-adjusted ≥ .774) were also non-significant. Left-right canal-pair comparisons were Benjamini-Hochberg corrected within each metric (six TC and six MSPEV tests). No left-right TC comparison was significant (all p-adjusted ≥ .717). For MSPEV, posterior canal responses exceeded anterior canal responses in the LARP pair during per-rotation (RP > LA; V = 26, p-adjusted = .021, r = -0.73) and in the RALP pair during post-rotation (LP > RA; V = 19, p-adjusted = .021, r = -0.75); the other two vertical MSPEV comparisons showed the same direction without surviving correction (LARP post-rotation: p-adjusted = .076; RALP per-rotation: p-adjusted = .079), consistent with higher posterior than anterior canal responses.

**Table 3.** Time constants (TC; seconds) and maximum slow-phase eye velocity (MSPEV; degrees/second) measured during per-rotation and post-rotation velocity step testing for each stimulated semicircular canal.

| Stimulated SCC | Per-Rotation |  | Post-Rotation |  | All |  |
|---|---|---|---|---|---|---|
|  | TC (s) | MSPEV (<sup>0</sup>/s) | TC (s) | MSPEV (<sup>0</sup>/s) | TC (s) | MSPEV (<sup>0</sup>/s) |
| **Left Horizontal** | 11.27 ± 4.85 | 60.08 ± 17.68 | 14.25 ± 4.61 | 70.40 ± 14.55 | 12.70 ± 4.92 | 65.04 ± 16.91 |
| **Right Horizontal** | 13.73 ± 6.31 | 62.66 ± 18.66 | 14.17 ± 6.69 | 65.90 ± 18.83 | 13.95 ± 6.44 | 64.31 ± 18.63 |
| 5<sup>th</sup> percentile | 4.59 | 33.41 | 4.56 | 38.25 | 4.47 | 33.69 |
| 95<sup>th</sup> percentile | 20.73 | 91.73 | 21.67 | 90.06 | 21.21 | 91.59 |
| **Left Anterior** | 5.21 ± 2.59 | 29.57 ± 8.62 | 5.97 ± 4.03 | 31.20 ± 13.09 | 5.58 ± 3.35 | 30.37 ± 10.92 |
| **Right Posterior** | 4.86 ± 2.36 | 41.22 ± 12.75 | 5.45 ± 2.05 | 41.58 ± 15.95 | 5.16 ± 2.20 | 41.40 ± 14.26 |
| 5<sup>th</sup> percentile | 1.28 | 18.75 | 1.77 | 13.44 | 1.28 | 15.00 |
| 95<sup>th</sup> percentile | 9.19 | 54.69 | 9.94 | 54.19 | 9.32 | 54.69 |
| **Right Anterior** | 5.15 ± 2.27 | 28.71 ± 10.82 | 6.75 ± 4.01 | 29.87 ± 12.68 | 5.99 ± 3.36 | 29.32 ± 11.69 |
| **Left Posterior** | 5.64 ± 2.46 | 39.19 ± 15.07 | 5.04 ± 2.55 | 36.91 ± 10.47 | 5.36 ± 2.49 | 38.10 ± 12.98 |
| 5<sup>th</sup> percentile | 1.62 | 15.50 | 2.04 | 15.66 | 1.72 | 15.44 |
| 95<sup>th</sup> percentile | 9.07 | 60.12 | 11.17 | 51.28 | 10.82 | 59.47 |

Values are presented as mean ± SD. Per-rotation values were obtained during constant-velocity chair rotation at 100<sup>0</sup>/s; post-rotation values were obtained following abrupt deceleration to rest. The “All” columns present the grand mean across per- and post-rotation conditions for reference. Individual semicircular canals are labeled by the excited canal for each rotation direction (e.g., LA = left anterior, RP = right posterior). TC = time for VOR slow-phase eye velocity to decay to 37% of its peak value; MSPEV = maximum slow-phase eye velocity observed during the response. LARP = left anterior-right posterior; RALP = right anterior-left posterior.

Preliminary normative upper limits of canal pair asymmetry were also computed for MSPEV (**Table 3**). Horizontal MSPEV asymmetry was lowest (per-rotation: 9.8 ± 7.7%, 95th percentile = 22.6%; post-rotation: 10.3 ± 10.6%, 95th percentile = 31.7%), while vertical canal pairs showed greater asymmetry (LARP per: 19.5 ± 14.2%, 95th = 39.0%; RALP per: 23.3 ± 17.4%, 95th = 51.3%). TC asymmetry was more variable than MSPEV asymmetry in five of six canal-pair conditions (e.g., LARP per-rotation TC: 29.2 ± 21.4% vs. MSPEV: 19.5 ± 14.2%); RALP per-rotation was the exception (TC: 16.2 ± 12.2% vs. MSPEV: 23.3 ± 17.4%). Both TC and MSPEV asymmetry may serve as lateralization indices for the vertical canals; however, the generally wider normative range for TC asymmetry suggests that MSPEV-based ARs may offer greater diagnostic precision, pending confirmation with future test-retest reliability data. The horizontal MSPEV asymmetry 95th percentile during per-rotation (22.6%) is slightly above the range of established horizontal caloric and rotational asymmetry norms (typically <20%) ^27, 28^; the wider post-rotation 95th percentile (31.7%) likely reflects the smaller effective sample and higher inter-subject variability and warrants confirmation in larger cohorts. The vertical canal values represent the first such reference data to our knowledge for vertical rotational testing.

ARs were computed for per- and post-rotation TC for each canal pair. Mean per-rotation AR values were: Horizontal 19.9 ± 15.8%, LARP 29.2 ± 21.4%, and RALP 16.2 ± 12.2%. Post-rotation AR values were: Horizontal 25.8 ± 23.6%, LARP 24.8 ± 19.7%, and RALP 26.1 ± 20.7%. Spearman correlations between per-rotation AR and vHIT gain asymmetry were non-significant for all canal planes (Horizontal: ρₛ = -0.023, p-adjusted = .951; LARP: ρₛ = -0.015, p-adjusted = .951; RALP: ρₛ = 0.029, p-adjusted = .951), indicating that mechanical canal gain asymmetry measured by VST does not closely track vHIT gain asymmetry in healthy subjects. Spearman rank correlations between VST time-constant AR and vHIT gain asymmetry were non-significant across all canal planes in both per-rotation (all |rs| ≤ 0.03, all padj ≥ 0.95) and post-rotation segments (all |rs| ≤ 0.45, all padj ≥ 0.43; Benjamini-Hochberg corrected across 6 tests), demonstrating that rotational-chair TC asymmetry does not track vHIT gain asymmetry in healthy subjects.

#### 3.2.1. Velocity storage analysis

Mean VSI values were +0.086 (Horizontal), +0.037 (LARP), and +0.014 (RALP). Although Horizontal VSI was numerically largest, consistent with the known dominance of velocity storage in the horizontal canal, a Friedman test revealed no significant difference across planes (χ²(2) = 0.32, p = .854), and pairwise Wilcoxon signed-rank tests confirmed no between-plane differences (Horizontal vs. LARP: V = 95, p-adjusted = 1.000; Horizontal vs. RALP: V = 118, p-adjusted = .548; LARP vs. RALP: V = 119, p-adjusted = .548, all Benjamini-Hochberg-corrected).

#### 3.2.2. Comparison of per- and post-rotation vVST parameters

Friedman tests revealed highly significant differences across the three canal planes for per-rotation TC (χ²(2) = 21.70, p < .001, Kendall’s W = 0.542) and post-rotation TC (χ²(2) = 26.95, p < .001, W = 0.642). Similarly, MSPEV differed significantly across planes during per-rotation (χ²(2) = 24.40, p < .001, W = 0.610) and post-rotation (χ²(2) = 25.81, p < .001, W = 0.615).

Following post-hoc comparisons (Benjamini-Hochberg-corrected pairwise Wilcoxon signed-rank tests) in the per-rotation condition, we found that TC in the horizontal plane was significantly higher than LARP (V = 208, p-adjusted < .001, r = 0.98) and RALP (V = 203, p-adjusted < .001, r = 0.93), with no significant difference between LARP and RALP (V = 99, p-adjusted = .837, r = -0.06). An analogous pattern was seen for MSPEV (horizontal > LARP: V = 206, p-adjusted < .001, r = 0.96; horizontal > RALP: V = 205, p-adjusted < .001, r = 0.95; LARP vs. RALP: V = 95, p-adjusted = .723, r = -0.10). Post-rotational analyses mirrored these results; horizontal TC and MSPEV were each significantly greater than their LARP and RALP counterparts (TC: horizontal vs. LARP: V = 230, p-adjusted < .001, r = 0.99; horizontal vs. RALP: V = 230, p-adjusted < .001, r = 0.99; LARP vs. RALP: V = 126, p-adjusted = .728, r = 0.09; MSPEV: horizontal vs. LARP: V = 227, p-adjusted < .001, r = 0.97; horizontal vs. RALP: V = 229, p-adjusted < .001, r = 0.98; LARP vs. RALP: V = 132, p-adjusted = .578, r = 0.14).

#### 3.2.3. Formal comparison with Morita et al. (2003)

To contextualize the present findings, we compared our vVST parameters directly with those reported by Morita et al. ^15^ the only prior study of vertical canal velocity step testing (**Table 4**). Morita et al. tested 10 healthy subjects using a 60⁰/s velocity step with the posterior SCC aligned in the earth-horizontal plane, whereas the present study used a 100⁰/s velocity step and tested both LARP and RALP canal planes. For horizontal TC, the present values (LH per 11.3 ± 4.8 s, LH post 14.3 ± 4.6 s; RH per 13.7 ± 6.3 s, RH post 14.2 ± 6.7 s) are somewhat shorter than those of Morita et al. (per 15.6 ± 5.1 s; post 16.2 ± 5.2 s). For vertical canal TC, the present values were close to Morita’s posterior canal estimates: LARP per 4.9-5.2 s and post 5.4-6.0 s (LA / RP); RALP per 5.1-5.6 s and post 5.0-6.7 s (LP / RA), versus Morita’s posterior canal values of 4.2 ± 1.5 s (per) and 5.7 ± 1.5 s (post). The small remaining differences may reflect the higher stimulus velocity (100 vs 60⁰/s), differences in desaccading, and Morita’s smaller sample (n = 10) and posterior-only configuration. When expressed as a proportion of stimulus velocity, our horizontal MSPEV ratios (per-rotation: 0.62; post-rotation: 0.68) align closely with Morita et al. (per: 0.70; post: 0.73). Our uncorrected vertical canal ratios (per: 0.35; post: 0.34-0.39) are somewhat lower than those reported by Morita et al. (per: 0.42; post: 0.50).

**Table 4.** Comparison of velocity step test (VST) parameters between the present study and Morita et al. (2003).

|  | **Present Study<br>(100<sup>0</sup>/s, n = 22–26)** |  | **Morita et al. 2003<br>(60<sup>0</sup>/s, n = 10)** |  |
|---|---|---|---|---|
| **Parameter** | **Per-Rotation** | **Post-Rotation** | **Per-Rotation** | **Post-Rotation** |
| **TC (s)** |  |  |  |  |
| Horizontal | 12.3 ± 4.8 | 14.0 ± 3.8 | 15.6 ± 5.1 | 16.2 ± 5.2 |
| LARP | 4.8 ± 1.9 | 5.7 ± 2.9 | -- | -- |
| RALP | 5.3 ± 2.3 | 5.6 ± 2.6 | -- | -- |
| Posterior (Morita) | -- | -- | 4.2 ± 1.5 | 5.7 ± 1.5 |
| **MSPEV (<sup>0</sup>/s)** |  |  |  |  |
| Horizontal | 62 ± 17 | 68 ± 16 | 42 ± NR | 44 ± NR |
| LARP | 35 ± 9 | 39 ± 15 | -- | -- |
| RALP | 35 ± 12 | 34 ± 10 | -- | -- |
| Posterior (Morita) | -- | -- | 25 ± NR | 30 ± NR |
| **MSPEV / Stimulus Velocity** |  |  |  |  |
| Horizontal | 0.62 | 0.68 | 0.70 | 0.73 |
| Vertical | 0.35 | 0.34-0.39 | 0.42 | 0.50 |

*NR = not reported (Morita et al. reported MSPEV as ratios to stimulus velocity; raw MSPEV values were derived by multiplying the reported ratio by stimulus velocity [60<sup>0</sup>/s]). Present study vertical MSPEV values are the mean of the two canals within each plane (e.g., LARP = mean of LA and RP). Morita et al. tested only the posterior canal. Dashes (--) indicate parameter not applicable or not tested in that study.*

Time constants (TC; seconds) and maximum slow-phase eye velocity (MSPEV; ⁰/s) are presented as mean ± SD for horizontal and vertical canal planes. The present study used a 100⁰/s stimulus velocity with both LARP and RALP canal plane positioning (plane-level means computed by averaging within subject across each plane’s two canals; restricted to participants with both per- and post-rotation values: horizontal n = 26, LARP and RALP n = 22); Morita et al. used a 60⁰/s stimulus velocity with posterior canal positioning only (n = 10). MSPEV/stim expresses MSPEV as a proportion of stimulus velocity to facilitate cross-study comparison. LARP = left anterior-right posterior; RALP = right anterior-left posterior.

#### 3.2.4. The correlation between VST and vHIT

There were no catch-up saccades defined as abnormal and the mean gain was normal for all SCCs in vHIT. Correlation coefficients were generally weak (|ρₛ| ≤ 0.39), with most ≤ 0.27. Spearman correlations between VST parameters and vHIT gain were computed for each of the six SCCs (24 comparisons). No correlation survived Benjamini-Hochberg correction. The strongest observed association was between right posterior post-rotation TC and vHIT gain (ρₛ = 0.394, uncorrected p = .095), which did not reach significance after Benjamini-Hochberg correction (p-adjusted = .872) (**Table 5**).

**Table 5.** Video head impulse test (vHIT) gain and Spearman rank correlations between vHIT gain and velocity step test (VST) parameters for each semicircular canal.

| Canal | vHIT Gain | Per-Rotation | Post-Rotation | Per-Rotation | Post-Rotation |
|---|---|---|---|---|---|
| Horizontal Right | 1.0 ± 0.1 | ρ<sub>s</sub> = 0.226 (p = 0.324) | ρ<sub>s</sub> = 0.226 (p = 0.313) | ρ<sub>s</sub> = 0.272 (p = 0.232) | ρ<sub>s</sub> = -0.086 (p = 0.702) |
| Horizontal Left | 0.92 ± 0.1 | ρ<sub>s</sub> = -0.068 (p = 0.757) | ρ<sub>s</sub> = -0.138 (p = 0.541) | ρ<sub>s</sub> = 0.343 (p = 0.109) | ρ<sub>s</sub> = 0.162 (p = 0.472) |
| Right Anterior | 0.82 ± 0.17 | ρ<sub>s</sub> = -0.155 (p = 0.553) | ρ<sub>s</sub> = 0.084 (p = 0.741) | ρ<sub>s</sub> = 0.091 (p = 0.728) | ρ<sub>s</sub> = 0.059 (p = 0.816) |
| Left Posterior | 0.7 ± 0.19 | ρ<sub>s</sub> = -0.237 (p = 0.301) | ρ<sub>s</sub> = -0.149 (p = 0.542) | ρ<sub>s</sub> = 0.052 (p = 0.823) | ρ<sub>s</sub> = 0.074 (p = 0.764) |
| Right Posterior | 0.81 ± 0.14 | ρ<sub>s</sub> = 0.385 (p = 0.104) | ρ<sub>s</sub> = 0.394 (p = 0.095) | ρ<sub>s</sub> = -0.013 (p = 0.957) | ρ<sub>s</sub> = -0.136 (p = 0.578) |
| Left Anterior | 0.75 ± 0.09 | ρ<sub>s</sub> = -0.011 (p = 0.962) | ρ<sub>s</sub> = -0.005 (p = 0.983) | ρ<sub>s</sub> = 0.008 (p = 0.975) | ρ<sub>s</sub> = 0.045 (p = 0.855) |

ρ<sub>s</sub> : Spearman rank correlation coefficient.
vHIT gain values (mean ± SD) are presented in the left column. Correlations (ρ<sub>s</sub>) are shown between vHIT gain and per-rotation TC, post-rotation TC, per-rotation MSPEV, and post-rotation MSPEV for each canal. Twenty-four correlations were tested (6 canals x 4 VST parameters); no correlation survived Benjamini-Hochberg correction for multiple comparisons.  vHIT was performed using the ICS Impulse system (Otometrics, Denmark); gain was calculated as the ratio of cumulative eye velocity to cumulative head velocity. TC = time constant; MSPEV = maximum slow-phase eye velocity; LA = left anterior; RA = right anterior; LP = left posterior; RP = right posterior.

#### 3.2.5. Transfer function analysis

Transfer function fitting was attempted on each subject’s SHA gain-frequency curve. However, the fits proved fundamentally under-constrained for our protocol: the physiologic VOR time constants (horizontal ∼12-14 s, vertical ∼5-6 s) correspond to corner frequencies of approximately 0.011-0.013 Hz (horizontal) and 0.028-0.033 Hz (vertical), both well below our lowest test frequency (0.08 Hz). Our three test frequencies therefore lie entirely on the asymptotically-flat upper portion of the high-pass curve, where the gain-frequency relationship is approximately constant and the time-constant parameter is non-identifiable. Across our cohort the gain varied minimally with frequency (e.g., LARP: 0.41, 0.37, 0.43 at 0.08, 0.16, 0.64 Hz respectively), and the majority of fits either failed to converge or returned implausibly short TCs that could not be reconciled with the directly-measured VST values. We therefore do not report SHA-derived time constants. This is a limitation of the test frequencies sampled rather than of the transfer-function approach itself, and the implications for protocol design are discussed below.

## Discussion

To our knowledge, this study is the first to characterize vertical SCC function using both vSHA and vVST rotational chair paradigms delivered via rotational chair. Together, these protocols extend established horizontal rotational testing into the LARP and RALP planes, providing a framework for comprehensive assessment of VOR dynamics across the low-to-mid frequency range most relevant to daily function ^1, 2, 24^.

The shorter time constants observed in vertical relative to horizontal planes align with a smaller velocity-storage contribution to the vertical VOR, a pattern long inferred from horizontal-plane physiology in normal and lesioned subjects ^29^. Velocity storage prolongs the VOR response beyond the mechanical time constant of the cupula and operates predominantly in the yaw plane ^25, 29, 30^. Accordingly, absolute vertical TC values in our sample (∼5-7 s across canals) were close to the published cupular time constant of ∼5 s, whereas horizontal values (∼11-14 s) were more than twice as long. The greater vertical phase lead at 0.08 and 0.16 Hz is consistent with this interpretation, because a shorter time constant produces more phase lead at low stimulus frequencies. Although the formal cross-plane comparison of velocity storage indicies in our sample (Friedman p = .854) was non-significant – likely reflecting insufficient power at n = 19 rather than a true absence of effect – the mechanistic interpretation remains well grounded primarily in the absolute TC values and wider literature.

Several methodological considerations must be noted. The present design cannot isolate central velocity storage mechanisms from concurrent differences in otolith and somatosensory input arising from the 70° head-tilt configuration. Furtheremore, vertical canal stimulation inherently generates a three-dimensional input comprising both vertical and torsional components; because torsional eye movements were not recorded, the contribution of this three-dimensional signal decomposition on observed TC differences remain undetermined. Nevertheless, the H-V TC ratio represents a promising candidate biomarker for disorders affecting the nodulo-uvular cerebellar complex ^17, 19, 31^. Formal validation in patients with documented central pathology along with study designs that decouple head orientation from canal stimulation will be necessary before clinical adoption.

Our findings broadly extend those of Morita et al.^15^, the only prior investigation of vertical canal velocity step testing. Horizontal TC values in our cohort were somewhat shorter than Morita’s but closely matched independent normative data from Yu et al. ^32^ (post-rotation TC 14.3–14.9 s at 90⁰/s, with TC defined as the time to 37% of peak). Vertical TC values closely matched Morita’s posterior canal estimates. Left-right symmetry was preserved for time constants (no pair differed after correction), whereas vertical canal pairs exhibited a consistent posterior-greater-than-anterior MSPEV asymmetry. These preliminary asymmetry reference ranges, particularly those derived from MSPEV, may prove useful in identifying unilateral vertical canal dysfunction, analogous to the us of caloric and horizontal VST asymmetry for lateralizing horizontal canal lesions ^7, 8, 33^.

The absence of significant correlations between rotational chair parameters and vHIT gain is both expected and clinically meaningful. These modalities assess fundamentally distinct regions of the VOR frequency spectrum: vHIT evaluates the high-frequency, high-acceleration response mediated primarily by direct three-neuron arc pathways ^3–5^, whereas SHA and VST characterize VOR dynamics at physiological frequencies where velocity storage and central integrative mechanisms predominate ^17, 24^. Consequently, a patient may present with normal vHIT gains yet exhibit marked abnormalities on rotational testing, or vice versa ^6, 19^. This dissociation underscores that vHIT and rotational testing provide complementary, non-redundant diagnostic information and should be combined for comprehensive vestibular assessment.

An important consideration for clinical SHA protocol design is the inclusion of sufficiently low stimulus frequencies. Standard test batteries typically begin at 0.01 or 0.02 Hz but these lowest frequencies are not always included. Our transfer function analysis demonstrated that the three frequencies tested here (0.08, 0.16, and 0.64 Hz) fall within the flat, mid-band region of the VOR transfer function – well above the corner frequency where phase lead increases and gain declines. This frequency selection accounts for the observed limitations in SHA-derived time constant estimation and highlights that calculating clinically meaningful time constant information from SHA requires inclusion of frequencies at or below 0.02 Hz ^12, 29^. Laboratories implementing vertical SHA testing should ensure their protocols sample this low-frequency region to capture the full dynamic profile.

Several study limitations warrant discussion. First, our cohort comprised young, healthy adults, limiting generalizability to older populations, in whom age-related declines in VOR gain and time constants are well documented ^26, 34, 35^. Second, while our sample size (n = 16-26 per cell) was adequate for initial normative estimates, it falls below the Clinical and Laboratory Standards Institute (CLSI)^36^ minimum (n = 120) for definitive reference intervals. The 5th-95th percentile bounds reported here should therefore be interpreted as exploratory. Third, test-retest reliability was not assessed, leaving the relative contributions of measurement error and true biological variation unknown. Multi-session reliability studies employing intraclass correlation coefficients and Bland-Altman analyses will be essential before these reference ranges can be applied clinically.

Beyond sample characteristics, several methodological constraints should be noted. Torsional eye movements were not recorded, and vertical canal stimulation in this paradigm required a 70° head tilt relative to gravity. This positioning simultaneously alters rotational axis orientation relative to the head, gravitoinertial otolith loading, and somatosensory input from the support surface—each of which can independently attenuate the apparent vertical VOR time constant. Indeed, horizontal tilted-head studies demonstrate gain reductions of 10–25% due to otolith-canal interactions ^25^. Furthermore, head alignment with the target canal plane was confirmed via visual inspection rather than quantitative tracking (e.g., IMU or optical verification). Given normal anatomical variation in canal plane orientation of up to 10°, minor off-axis alignment is inevitable, potentially distributing stimulation across adjacent planes and reducing apparent vertical gain. Finally, our eye-velocity calibration protocol used a combined VOR-plus-smooth-pursuit stimulus rather than an isolated VOR target, which may introduce a systematic scaling factor. Most importantly, the clinical sensitivity of these vertical paradigms remains to be established through studies in patients with known vestibular pathology, including unilateral and bilateral vestibulopathy ^37^, vestibular migraine ^8^, and central vestibular disorders.

## Conclusion

In summary, the vertical SHA and VST paradigms described here provide preliminary descriptive reference data for three-dimensional rotational VOR assessment using vSHA and vVST. The findings demonstrate that vertical paradigms provide non-redundant information relative to horizontal rotational testing and vHIT, with velocity storage-driven time constant differences carrying potential diagnostic value for central vestibular pathology. Clinical validation is the critical next step toward comprehensive VOR assessment.

## Acknowledgements

The authors thank Joseph Furman for his early contributions to the methodology and technical revisions to the manuscript.

## Author Contributions

FM and EW contributed equally as co-first authors. FM: conceptualization, methodology, investigation, data curation, formal analysis, writing - original draft, writing - review C editing. EW: conceptualization, methodology, investigation, data curation, formal analysis, writing - original draft, writing - review C editing. MH: investigation, data curation, writing - review C editing. BM: investigation. AK: resources, funding acquisition, data curation, writing - review C editing. MEH: conceptualization, supervision, writing - review C editing.

## Statements and Declarations

Alexander Kiderman is the VP Technology Development and is an employee and shareholder of Spryson America, Inc. Additionally, Alexander Kiderman holds a U.S. patent related to this work. All other authors declare that the research was conducted in the absence of any commercial or financial relationships that could be construed as a potential conflict of interest.

## Ethical approval and informed consent statements

This study was approved by the Institutional Review Board of The University of Miami (IRB#20190034). All procedures were conducted in accordance with the Declaration of Helsinki. Written informed consent was obtained from all participants prior to testing.

## Data availability statement

A de-identified dataset used in this study is available upon request to the corresponding author. Please include your name, affiliation, and email address in your request.

